# Is virtual care the new normal? Evidence supporting Covid-19’s durable transformation on healthcare delivery

**DOI:** 10.1101/2022.03.08.22272020

**Authors:** Soumik Mandal, Batia Wiesenfeld, Devin M. Mann, Katharine Lawrence, Rumi Chunara, Paul A. Testa, Oded Nov

## Abstract

**Objective:** Despite the surge of telemedicine use during the early stages of the coronavirus-19 (COVID-19) pandemic, research has not evaluated the extent to which the growth of telemedicine has been sustained during recurring pandemic waves. This study provides data on the long-term durability of video-based telemedicine visits and their impact on urgent and non-urgent healthcare delivery from one large health system in New York City.

**Materials and Methods:** Electronic health record (EHR) data of patients between January 1st, 2020 and November 30th, 2021 were used to conduct the analyses and longitudinal comparisons of telemedicine or in-person visit volumes. Patients’ diagnosis data were used to differentiate COVID-19 suspected visits from non-COVID-19 ones while comparing the visit types.

**Results:** While COVID-19 prompted an increase in telemedicine visits and a simultaneous decline in in-person clinic visits, telemedicine use has stabilized since then for both COVID-19 and non-COVID suspected visits. For COVID-19 suspected visits, utilization of virtual urgent care facilities is higher than the trend. The data further suggests that virtual healthcare delivery supplements, rather than replaces, in-person care.

**Discussion:** The COVID-19 pandemic has transformed the use of telemedicine as a means of healthcare delivery, and the data presented here suggests that this is an enduring transformation.

**Conclusion:** Telemedicine use increased with the surge of infection cases during the pandemic, but evidence suggests that it will persist after the pandemic, especially for younger patients, for both urgent and non-urgent care. These findings have implications for the healthcare delivery system, insurers and policymakers.

## INTRODUCTION

Before the COVID-19 pandemic, telemedicine’s growth in the US was incremental, and only 8% of Americans were reported to have used telemedicine in 2019 [1]. In prior studies on telemedicine adoption, limited reimbursement, lack of comfort with telemedicine technologies by patients and providers, and few compelling justifications outside of rural medicine for replacing in-person care were identified as the most significant barriers to wider adoption [2]. However, starting in March 2020, the telemedicine landscape in the US changed rapidly when the World Health Organization (WHO) declared coronavirus disease 2019 (COVID-19) a pandemic and a nationwide emergency was declared in the US^1^. Healthcare systems quickly turned to telemedicine solutions to help maintain existing operations while meeting the new demands imposed by rising COVID-19 cases. Studies report an exponential growth (683%) of telemedicine in the acute pandemic period (between March and April, 2020) [3]. Prior reports relating the growing prevalence of telemedicine to a steady decline in in-person clinic visit volumes suggested that telemedicine was at least partially replacing clinic visits. However, little research on the durability of this migration, particularly in the post-acute pandemic period, is available. This report, drawn from a large academic healthcare system in New York City (NYC), empirically demonstrates the persistence of both patients’ and providers’ migration to telemedicine during the recurring waves of the pandemic.

Prior to large-scale availability of any vaccine or effective therapies, social distancing and quarantine were the only widely-accepted approaches to minimizing viral spread, creating a compelling reason for finding alternatives to in-person care, often involving telemedicine [4]. To ensure the pace of scaling telemedicine capacity matched growing demand, implementation with rapid iterative improvements was preferred over perfect execution [3]. Where possible, existing technology and vendors were leveraged instead of investing limited time into procuring all new technology. As a result, in many cases, telemedicine infrastructure spanned multiple technologies and platforms, supported different modalities (voice-based over the telephone, or video-based) rather than any standardized implementation, and evolved rapidly in a short period of time, all of which might negatively affect patients’ satisfaction [5] and patient retention with telemedicine.

Since the first acute phase of the pandemic passed, breakthrough developments in vaccine research, vaccination, and treatment protocols have made partial to full resumption of in-person activities possible. Are patients still inclined to use telemedicine when in-person clinic visits are available?

To address this question, we use data from a large academic healthcare system in NYC with a robust telemedicine infrastructure to characterize patients’ visit types between January 1, 2020 and August 31, 2021. The long timeframe allows us to compare telemedicine visit volumes and in-person clinic visits during recurring waves of pandemic intensity. We use data regarding encounters, visits, diagnoses, and patients’ age to identify the age groups that accessed care through telehealth or otherwise during this period.

## METHODS

New York University Langone Health (NYULH), a large academic healthcare system in NYC, consists of 8,077 healthcare providers across 5 hospitals and 500+ ambulatory locations all connected on a single electronic healthcare record system (Epic, Verona, WI). To enable its virtual health services, NYULH uses a single instance of the Epic health record with 7,545,427 active patients leveraging an integrated video visit platform. Before COVID -19, NYULH implemented telemedicine capabilities in approximately 25 locations. Its largest investment was in virtual urgent care, a video visit experience tightly integrated into its enterprise EHR and its patient portal. Patients accessing virtual urgent care were able to check-in, pay, and have a video-enabled consultation with an NYULH emergency medicine physician that was reimbursed by multiple local payors. This system had been running since 2018 and prior to the pandemic had <100 visits on a typical day. The visits were managed by a pool of about 40 ED providers who took visits either on-site or at home. Other than the EHR integrated platform, NYULH also utilized Webex by Cisco^2^ and telephone calls for a very brief period for providing telehealth services (<1%).

Patients access the virtual services through the NYULH app built upon the Epic MyChart suite of patient tools and using standard APIs made available by Epic. In addition, NYULH has deployed native open scheduling technologies as well as custom features enabling simplified virtual care access and matriculation. Patient satisfaction and engagement is assessed via brief text message surveys at the close of telemedicine encounters leveraging Q-Reviews (NY, NY). The questions in the survey assess domains including satisfaction with the visit, likelihood to use it again and likelihood to refer a friend to the service. The mean of the responses to these three questions is reported in this paper as the satisfaction rating. Data for this survey were queried from the hospital EHR system from January 1^st^, 2020 to November, 2021.

We examined the diagnosis information from patients’ visits to evaluate whether telehealth utilization was skewed towards COVID-19 suspected visits, which may evaporate in the post-pandemic world. COVID-19 suspected visits were identified from diagnosis codes containing relevant respiratory issues via partial matching with the following 23 keywords [4, 7]: 1) COVID, 2) RESPIRATORY DISTRESS, 3) FLU, 4) URI, 5) PNEUMONIA, 6) FEVER, 7) SHORTNESS OF BREATH, 8) COUGH, 9) DYSPNEA, 10) PHARYNGITIS, 11) BRONCHITIS, 12) SINUSITIS, 13) CHEST PAIN, 14) MUSCLE PAIN, 15) HEADACHE, 16) SORE THROAT, 17) CONGESTION, 18) TACHYPNEA, 19) MALAISE, 20) LUNG INFILTRATES, 21) HYPOXIA, 22) OPACITIES, and 23) ARDS. Additional data on total COVID-19 cases per day in NYC was used to compare the patients’ visit types and volumes with confirmed COVID-19 cases in the city^3^. Telemedicine encounters were then categorized into two groups: 1) virtual urgent care (VUC) and 2) non-urgent telehealth or ambulatory care visits. Descriptive statistics were computed to estimate rates of telemedicine visits in urgent care and non-urgent care settings, both COVID-19 suspected and not.

To study whether virtual healthcare delivery supplements or replaces in-person care while controlling for seasonal differences, we calculated the average per-patient in-person and virtual visits in two periods: pre-pandemic (June 2019 to November 2019) and post-acute pandemic (June 2021 to November 2021.

## RESULTS

During the 23-month window, a total of 2,475,278 telehealth visits were recorded, which is nearly one-third the volume of all in-person visits recorded (7,930,049) in the same time frame. Nearly 90% of all video visits (2,203,756) were for non-urgent care, and the remaining 10% (251,006 visits) were for urgent care. The telehealth volumes were insignificant (<100 per day) before the COVID-19 pandemic (January and February, 2020), and peaked during the month of April, 2020 (240,904 visits), during the acute pandemic period. The longitudinal distributions of visit type percentages (see Figure 1) suggest that each increase in the volume of urgent or non-urgent telehealth visits coincided with a simultaneous decline of in-person visits. This was particularly evident during the acute pandemic period (between March, 2020 and April, 2020) and also during the following winter season (between October, 2020 and January, 2021). Overall, the distributions suggest that volumes of telehealth visits peaked in the acute pandemic phase, and stabilized since then at a rate much higher than before the pandemic.

**Figure 1:**
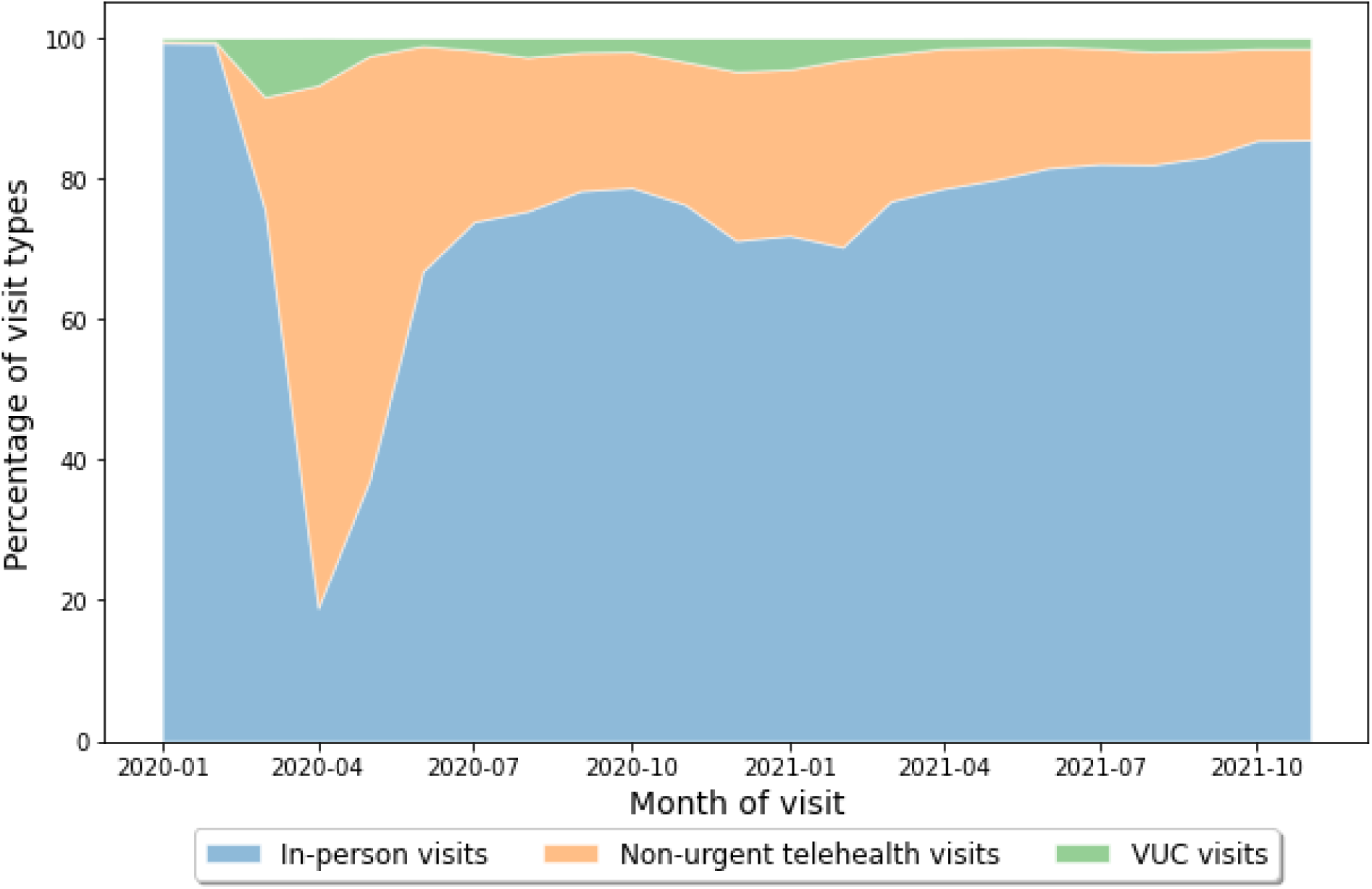
Share of visits in telemedicine urgent care (VUC), non-urgent telehealth care (non-VUC), and in-person care. X-axis represents the month of the visits.

Of all the visits (in-person and telemedicine visits combined) recorded in this period, 770,372 cases (∼7.4%) reported at least one of the COVID-19 symptoms. Among these COVID-19 suspected cases, 511,172 (66.35%) were recorded in in-person facilities, and the remaining 259,200 (33.65%) were telehealth visits. Thus, the ratio of virtual and in-person visit volumes for COVID-19 suspected cases is similar to the ratio for all recorded visits.

The distribution of video visit types across urgent and non-urgent care shows greater utilization of urgent care services for COVID-19 symptoms than what was witnessed in general (see Figure 2): the virtual visit volumes for COVID-19 suspected cases were far more evenly distributed between VUC and non-urgent facilities than what was witnessed for all recorded visits. About 48.08% of COVID-19 suspected visits (124,632) were for urgent care, whereas non-urgent telehealth visits constituted the remaining 51.92% cases (134,568).

**Figure 2:**
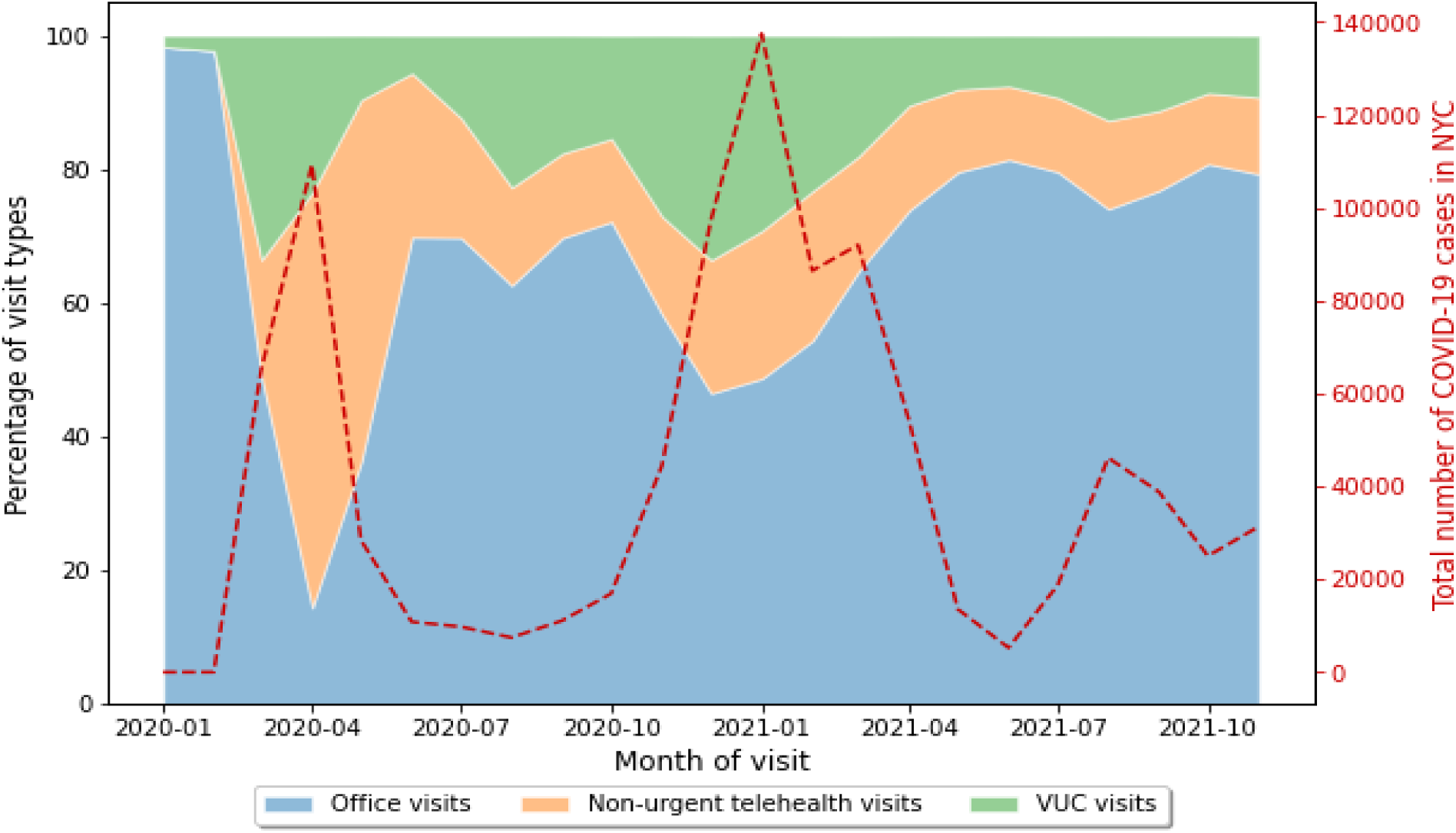
COVID-19 suspected visit type percentages and confirmed COVID-19 cases in NYC between January, 2020 and November, 2021

The distributions of COVID-19 suspected visit types were further compared with confirmed COVID-19 cases in NYC [8] in the same period (see Figure 2). The distributions suggest that increases in COVID-19 cases coincided with increased telehealth visits, especially to urgent care facilities, and at the same time decreased in-person or office visits. This is particularly evident in both the first (March and April 2020) and second waves (November, 2020 and February, 2021) of the pandemic, when COVID-19 cases spiked in NYC. Overall, the fraction of telehealth visits for urgent care changed more dynamically than non-urgent care, and showed strong correlation (Pearson’s coeff: 0.721) with COVID-19 numbers in NYC.

To examine the utilization of telemedicine outside of the immediate COVID needs, we analyzed visit types for non-COVID suspected cases (9,634,955 visits) separately (see Figure 3). Since non-COVID suspected visits accounted for almost 92.6% of all visits recorded, their distributions across visit types were near-identical to those of all visits. Overall, these distributions suggest that although COVID-19 prompted rapid scaling and utilization of telemedicine, its use grew and then stabilized at a higher level for non-COVID suspected cases as well.

**Figure 3:**
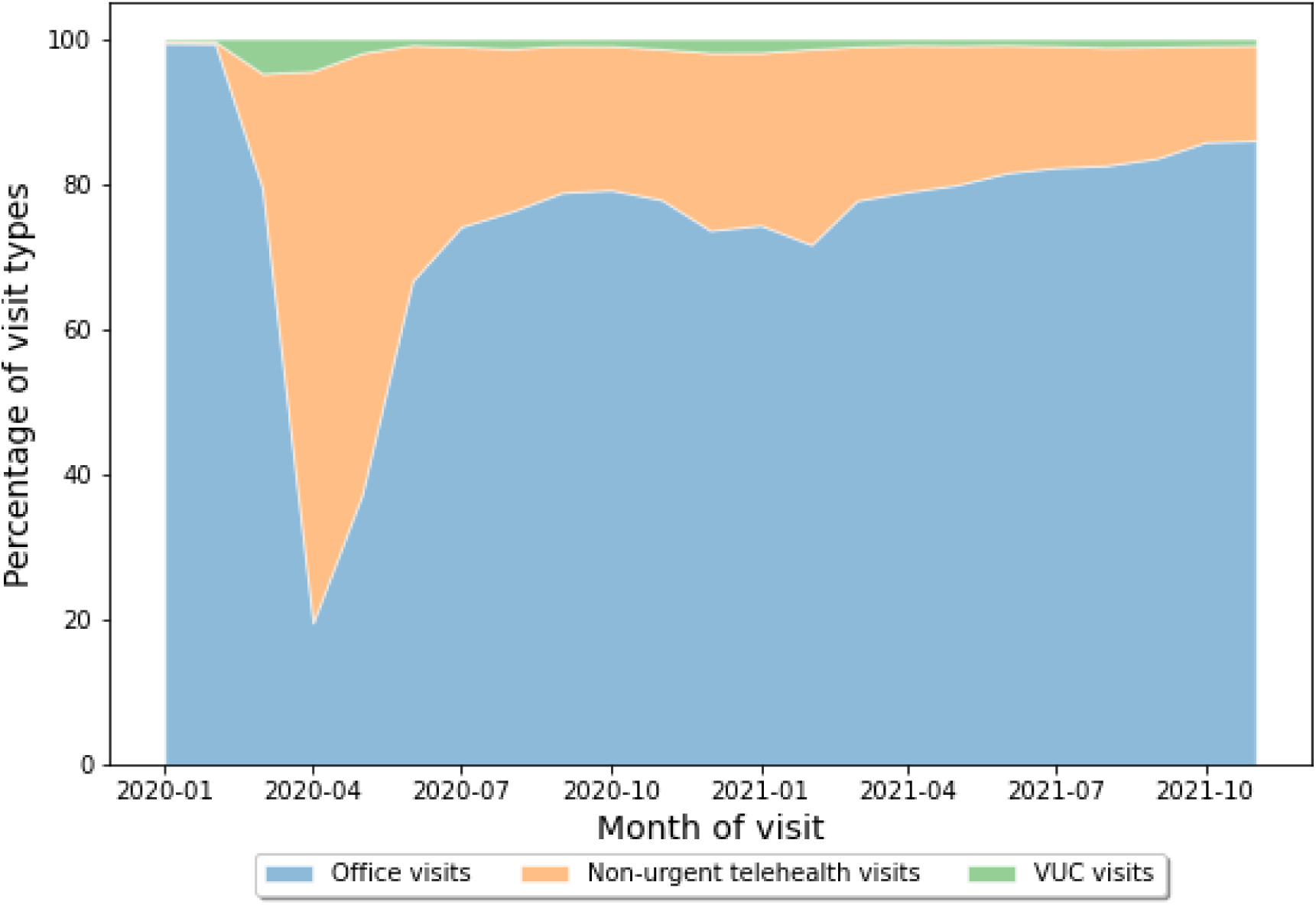
visit types and their percentages for non-COVID suspected cases

Prior studies that analyzed demographics of patients during the initial surge in use of telemedicine reported telemedicine utilization as highest among young patients [4, 6]. We also evaluated the age group of patients in our data to examine any changes in patient demographics throughout this period. For each telemedicine visit record, we determined patient’s age at the time of the visit to identify which age groups were more dominant users of telehealth (see Table 1). Regardless of virtual visit type, the 20-39 year old age group accounted for the largest proportion of telemedicine visits. Additionally, compared to ambulatory visits, the distribution of telehealth visits in urgent care was further skewed among young patients. A one-way ANOVA of visit volumes by age group was found to be significantly different for all telehealth visits combined and for VUC visits and non-VUC visits separately (*p-*value < 0.005*** in all three cases).

**Table 1:** Distribution of telemedicine visits by age.

|  |
| --- |
| Telemedicine visits by age |

| Age group (Yrs) | Urgent Care (%) | Non-urgent care (%) | All telemedicine care (%) |
| --- | --- | --- | --- |
| <20 | 4.83 | 12.66 | 12.02 |
| 20-39 | 66.27 | 35.78 | 38.81 |
| 40-59 | 22.58 | 29.38 | 28.65 |
| 60-79 | 5.88 | 19.55 | 18.13 |
| >=80 | 0.44 | 2.63 | 2.39 |

Figure 4 illustrates the trends in virtual health visits among different age groups throughout the entire period considered in this study. The figure shows that while the largest contributing age group’s telehealth use was steady throughout this period, the other age groups’ utilization of telehealth grew each time the number of COVID-19 cases surged causing a drop in percentage of telehealth utilization for the largest contributor group. Overall, the visit volumes in Table 1 and the percentages in figure 4 suggest that older patients (80 or above years) remained less likely (about 2%) to use telemedicine service.

**Figure 4:**
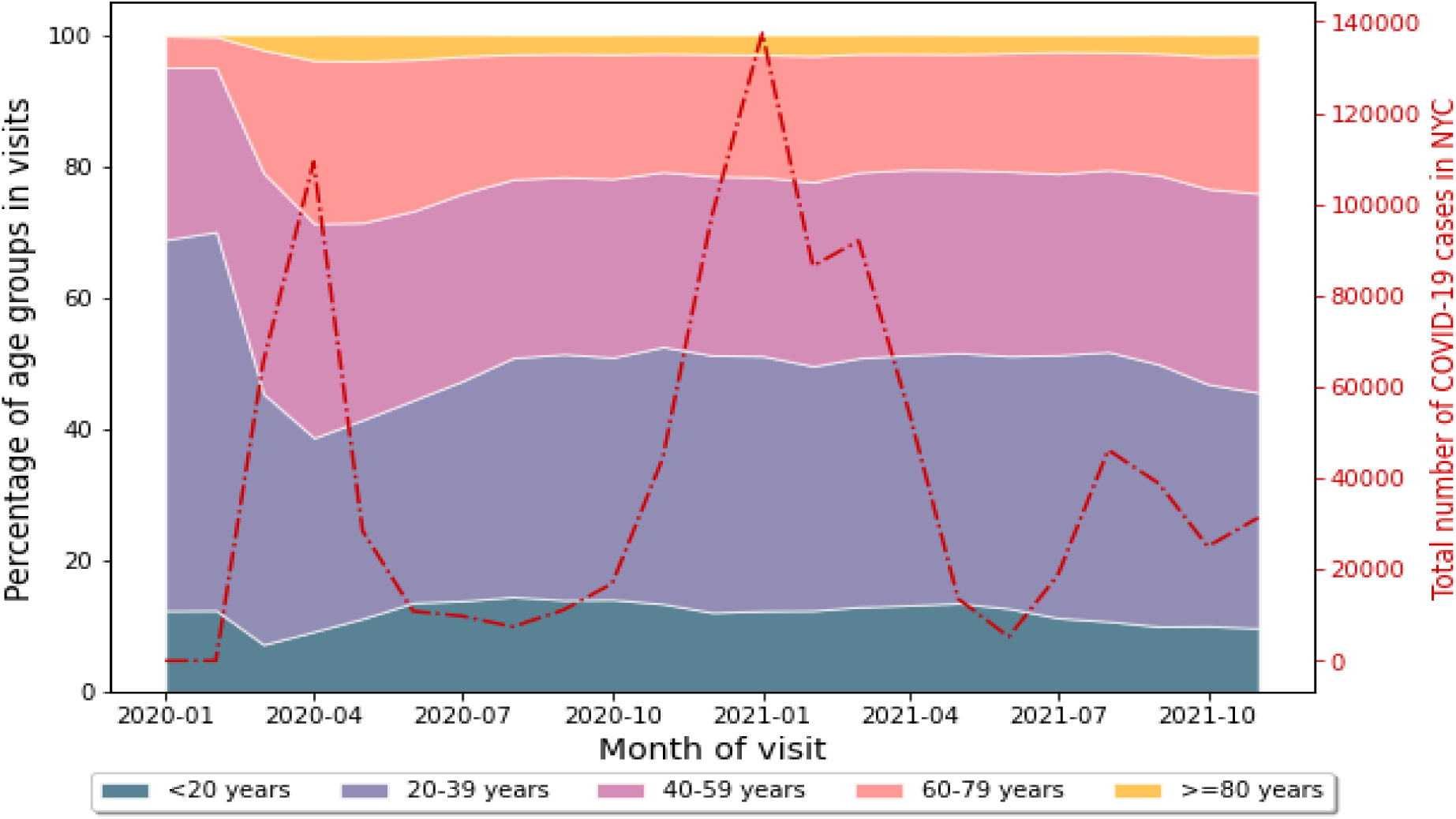
Stacked area graph of trends in age groups’ use of telemedicine

Despite the inexperience of providers rapidly adopting telemedicine, patients’ satisfaction with telemedicine visits remained unchanged during the acute pandemic phase (pre-COVID: N: 847, mean satisfaction: 4.38/5; acute-COVID: N: 1693, mean satisfaction: 4.38/5). While the number of telehealth visits stabilized since then, patients’ satisfaction with virtual urgent care has steadily increased (for 23 months: N: 11,878, mean satisfaction: 4.55/5), despite 16.5% reporting cases of technical issues with virtual urgent care visits. More than 70% of survey respondents felt they saved at least an hour of time (including travel time) by using virtual urgent care services.

Finally, patients’ average video visits increased from 0.013 in the pre-pandemic period (June 2019 to November 2019) to 0.588 between June 2021 to November 2021. During the same time frames, the average number of in-person visits had slightly declined from 2.928 to 2.894 per patient.

## DISCUSSION

This report demonstrates the transformation of telemedicine-based healthcare in NYC, the epicenter of the early stage of the COVID-19 pandemic in the US. The results demonstrate that while the pandemic catalyzed the rapid growth in telemedicine utilization, the migration from in-person emergency department visits to telemedicine has stabilized and has sustained, with a new mix of healthcare delivery in which telemedicine utilization is largely resilient to fluctuations in COVID-19 cases. Trends in telemedicine visit volumes for cases that are not COVID-19 related reinforce the position of telemedicine as a new norm in healthcare delivery. With an increase in virtual urgent care use and a simultaneous decrease in emergency room visits at each recurring pandemic wave, these data substantiate the critical role of telemedicine in expanding emergency room capacity during a pandemic, providing a bulwark to slow the infection rates.

In the non-urgent care setting, the transition to telemedicine suggests a delayed, but more pronounced increase compared to urgent care cases. While urgent care video visits are expected to rise in more acute settings, telemedicine visits for non-urgent cases have become part of the norm in healthcare access and accelerated telemedicine utilization. In all, while the urgent care visits opened the door for wider adoption of telemedicine during the pandemic, it is non-urgent video visits that are driving the sustainability of telemedicine adoption.

The longitudinal analysis of telemedicine patients’ demographics suggests that younger patients are likely to remain over-represented in telemedicine use. Compared to other age groups, the low rates of telemedicine adoption among oldest adults is likely due to preference for emergency department visits, lower rates of technology adoption [9], and other reasons. While recent reports suggest that smartphone adoption and internet use have more than doubled in the last seven years among older adults [10], there remains a notable digital divide between younger and older Americans. If increase in technology adoption can transpire into telemedicine use uptake among seniors remains to be seen.

Furthermore, an 18% increase of combined telehealth and in-person visits was observed from pre-COVID era (2019) to recent times (2021), and telehealth was responsible (106%) for this increase, which suggests that virtual-care delivery supplements, rather than displaces in-person care. This may be a result of the enhanced access to care that virtual care provides allowing people with geographic, logistic or other barriers to in-person care to more regularly access care. Virtual care may be unlocking unmet needs of underserved patient populations and potentially improve health equity and reduce health disparities if made accessible to inclusive populations.

While prior studies have found evidence of telemedicine access disparities reflect those in in-person healthcare access [6], whether telemedicine access disparities have reduced over time through increased and sustained use, remains to be investigated. Nonetheless, evidence suggests healthcare organizations need to allocate additional resources to virtual care, which should not come at the expense of in person care. For providers, the transition means quickly developing and adjusting skills in virtual rapport building, empathy, diagnosis, and counseling.

The trends in telehealth visits suggest proportionately larger role for urgent care facilities, particularly for COVID-19 care, compared to non-urgent and non-COVID care. With the emergence of new Covid variants [11], and the strong correlation between virtual urgent care visits and COVID-19 confirmed cases, demand for virtual urgent care is not expected to decrease in the near future. More importantly, our observation of a new pattern of sustained demand for non-urgent, non-Covid related virtual care has enormous implications for healthcare delivery and equity. For patients, the steady increase of satisfaction with virtual visits indicates their acceptance and willingness to persist with telehealth services in the future. Whether or not broad reimbursement of virtual visits will continue [12], will be one of the factors determining the future of telemedicine as a mainstream mode of healthcare delivery in the U.S.

## Data Availability

The data used in this study came from our EHR system, and can be shared in a limited basis upon reasonable request to the authors. COvid-19 city-specific data came from NYC Open data source and given below

https://www1.nyc.gov/site/doh/covid/covid-19-data.page

## ACKNOWLEDGEMENTS

The authors would like to thank (anonymized) for her thoughtful review and (anonymized) for her assistance in the manuscript preparation.

## CONTRIBUTORSHIP

All authors made substantial contributions to conception and design; acquisition of data or analysis and interpretation of data; were involved in drafting the manuscript or revising it critically for important intellectual content; gave final approval of the version published; and agreed to be accountable for all aspects of the work.

## Footnotes

1 https://www.cdc.gov/museum/timeline/covid19.html

2 https://www.webex.com/

3 https://www1.nyc.gov/site/doh/covid/covid-19-data.page

